# COMPARATIVE ANALYSIS OF COVID-19 MORTALITY IN BRAZIL, RIO DE JANEIRO, CAMPOS DOS GOYTACAZES, MACAÉ, CABO FRIO AND RIO DAS OSTRAS^1^

**DOI:** 10.1101/2020.09.17.20196444

**Authors:** Antonio C. C. Guimarães, Karla Santa Cruz Coelho, Kathleen Tereza da Cruz, Bárbara Soares de Oliveira Souza, Janimayri Forastieri de Almeida, Gustavo Fialho Coelho, Gabriella Ramos Lacerda Ferreira

**Author notes:** A Portuguese version of this work can be found here: https://drive.google.com/file/d/1VwWD02Hsn9DQIsi-RHwxSEKppiykhCLn/view?usp=sharing.

## Abstract

**Objective:** To analyze quantitatively and comparatively the deaths by COVID-19 of the four largest municipalities in the North of Rio de Janeiro and Baixada Litorânea of Rio de Janeiro, within the national context.

**Methods:** We used data from the Civil Registry and demographic information to elaborate a general picture of the pandemic up to the 31st epidemiological week in several aspects: evolution, scope, age, sex, race and impact on other causes of death.

**Results:** We characterized the evolution of the pandemic. We found an exponential dependence on the mortality rate by age and a higher lethality in the male population. We determined that COVID-19 represents an important fraction of the causes of death in 2020, being associated with a significant excess of deaths in relation to 2019 and also with the change in mortality patterns due to other causes.

**Conclusion:** Mortality is an effective and powerful indicator for understanding the infection and its pandemic, and it must be taken into account during the construction of public policies to deal with it.

## 1. Introduction

The year 2020 will be marked by one of the biggest humanitarian crises in the world, caused by SARS-CoV-2, responsible for the pandemic of COVID-19. In Brazil, the pandemic began to be detected in February and claimed fatalities in March, having a profound impact on public health and the country’s social and economic activities^1,2^.

As of August 1^st^, 2020, Brazil registered 2,707,877 cases of COVID-19, with a total of 93,563 patients progressing to death. With these additions to the statistics, the country appears in the 9^th^ worldwide position of the incidence of the disease with a coefficient of 12,886 cases / million inhabitants and occupies the 10^th^ position in deaths due to the pandemic of the new coronavirus with a coefficient of 445 deaths / million inhabitants, according to the Ministry of Health (MS) ^3^. The high estimated underreporting of cases and its variability in time and location, due to policies and availability of testing, make the number of cases a problematic measure for the reliable monitoring of the pandemic in the country^4^. The number of deaths, although it can also suffer from problems, seems to be a more accurate and reliable indicator^5^.

Since the 1970s, Brazil has been adopting measures to systematically register all deaths occurring in the national territory with the aim of taking each death as an epidemiological event. Death starts to have a name, address and cause, it is investigated by health authorities and of public health interest, constituting a key element in the diagnosis of the health situation of a given territory^6^. Several databases record deaths in the country, attributing cause of death to them. These records are made by different professionals, following different standards (which change over time), so there is not always a total agreement between the various databases. Some databases have a more definitive registration character, such as the Civil Registry (RC)^7^. Others, such as the Mortality Information System (SIM), may undergo revisions and changes to the record of old deaths after conducting the epidemiological investigation, but are not publicly available at the national level in real time. From these databases it is possible to analyze the historical statistical series of available mortality, detect trends and raise hypotheses about the epidemiological behavior of each disease.

The present study uses mortality indicators to carry out an exploratory analysis of the pandemic COVID-19 in the four largest municipalities in Northern Fluminense (Campos dos Goytacazes and Macaé) and Baixada Litorânea (Cabo Frio and Rio das Ostras) in the State of Rio de Janeiro, placing them on the national scene, as well as contributing to the understanding of some universal aspects. The municipalities mentioned are regional hubs and have intense human exchange, in addition to sharing economic identity related to oil exploration, being the destination for workers from other regions of the country and even from other countries.

This research is carried out within the framework of the Multidisciplinary Working Group to Confront COVID-19, formed by researchers from the Federal University of Rio de Janeiro (Campus UFRJ-Macaé) and the Federal Fluminense University (UFF-Rio das Ostras)^8^.

## 2. Methods

This is a descriptive epidemiological research based on Civil Registry (CR) databases.7 It was considered the period of 20 weeks, from Mar/15/2020 to Aug/1/2020, from the first death registered nationally in the 12th to 31st epidemiological week (E.W.).

The CR database is fed by the registry offices with the information contained in the death certificates filled out by doctors who are legally responsible for them, and the registry is made in the municipality where the death occurred, regardless of the municipality of residence of the deceased. Currently, this database only allows to know the number of deaths that occurred in a given municipality and the number of non-residents of that municipality who died there. Another limitation is the delay between death and registration in the RC database, which is usually less than a week, but may take longer.

The compilation of data extracted from the RC was delimited by period and location (Brazil, State of Rio de Janeiro, Rio de Janeiro capital, Campos dos Goytacazes, Macaé, Cabo Frio and Rio das Ostras), and we organized and analyzed the data accordingly with variables of interest: causa mortis (“Cardiac causes” tab on the RC Portal, and we also looked at the same period in 2019 for comparison), place of death, age, sex and race/color.

We used demographic data from the Brazilian Institute of Geography and Statistics for various calculations and analyzes. For the mortality rate (death by population) the estimated total population for 2020 was used: Brazil 211,755,692, State of Rio de Janeiro 17,366,189, Rio de Janeiro capital 6,747,815, Campos dos Goytacazes 511,168, Macaé 261,501, Cabo Frio 230,378 and Rio das Ostras 155,193 inhabitants^9^. For calculations referring to population subgroups (age, sex and color), data from the 2010 census were used, without paying attention to possible corrections due to population growth since then^10^.

The software used to compile, process, analyze and generate data visualizations were: Google Spreadsheet, Language R, Power BI and Grace.

This study is part of the research project entitled “Coping with COVID-19 in the North Fluminense Region and Baixada Litorânea: Actions, perspectives and impacts”, approved by the Research Ethics Committee of the Federal University of Rio de Janeiro / Campus UFRJ-Macaé Professor Aloísio Teixeira, under nº CAAE: 32186520.7.0000.5699.

## 3. Results

We focus our analysis on CR data by location. Figure 1 places the study geographically, centered on the regions of Norte Fluminense and Baixada Litorânea, belonging to the State of Rio de Janeiro, Brazil, providing an overview of the reach of the COVID-19 pandemic in terms of its mortality coefficient.

**Figure 1.**
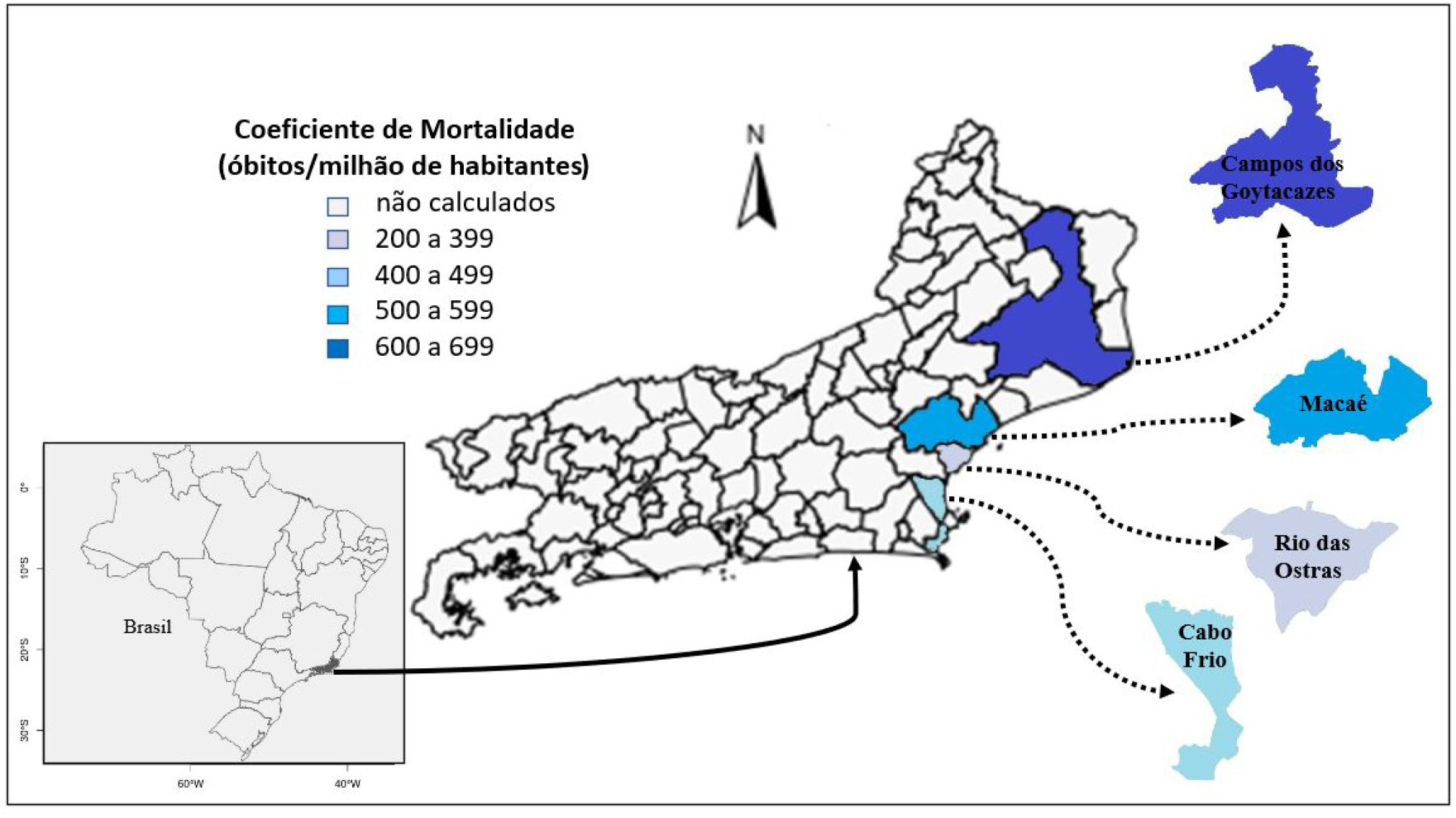
Locations studied: Brazil, State of Rio de Janeiro and municipalities of Campos dos Goytacazes, Macaé, Cabo Frio and Rio das Ostras. The 4 municipalities analyzed were colored on a scale indicative of the mortality coefficient for COVID-19 at the end of the 31^st^ S.E. (August 1^st^, 2020).

Figure 2 shows the evolution of mortality due to COVID-19 in the analyzed locations. The pattern developed by the country as a whole is one of rapid initial growth until it reaches a plateau around the 19^th^ E.W. that lasts until the end of the period considered (31^st^ W.E.). The state of Rio de Janeiro has experienced an extremely rapid and intense growth in deaths since the 12^th^ E.W., reaching a peak in the 19^th^ E.W., followed by a decline to a level close to the national average. The interior of the state (RJ interior) follows a similar pattern, but less intense, to that of the state, probably dominated by the municipalities of the metropolitan region that form a highly connected population group, as they are part of the same conurbation as the capital. The municipalities analyzed, farther from the metropolitan region, have a slower initial growth, with late acceleration in mortality, reaching maximum values only between the 22^nd^ E.W. (Cabo Frio) and 27^th^ E.W. (Campos dos Goytacazes). All municipalities have fluctuations in the weekly mortality rate and a consistent phase of decline is not characterized in any of them. The weekly mortality rate for all studied locations converges at the end of the period considered to be between 15 and 50 deaths per COVID-19 per million inhabitants per week, which also includes the national average (30 deaths / million / week).

**Figure 2.**
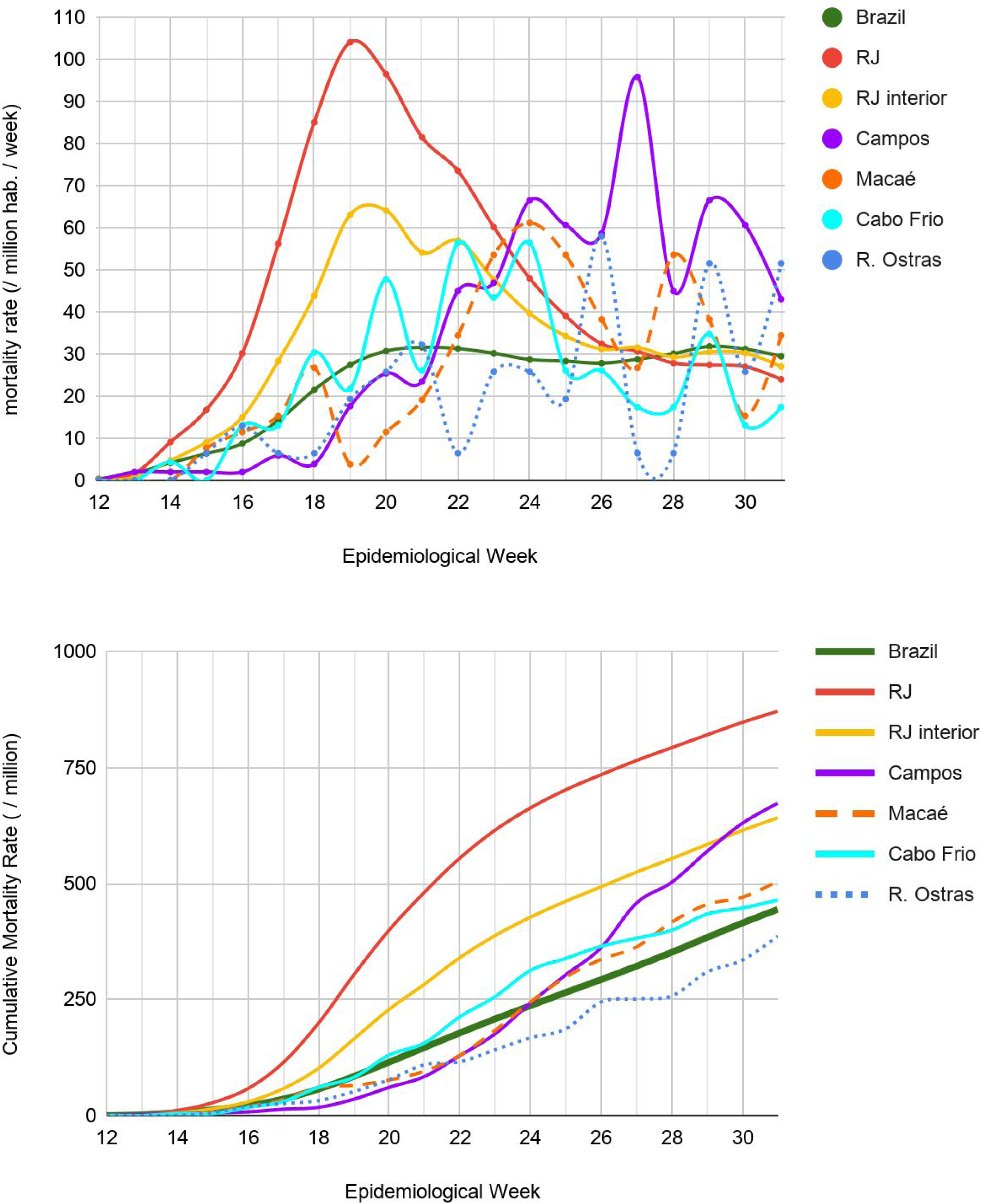
Evolution of the number of deaths by COVID-19 per epidemiological week for: Brazil, RJ (state and interior), Campos dos Goytacazes, Macaé, Cabo Frio and Rio das Ostras. Top panel: weekly rate; lower panel: accumulated mortality coefficient.

The cumulative number of deaths by the respective populations (figure 2, lower panel) clearly shows that the epidemic continued to grow in all the locations analyzed at the end of the considered period, from the municipal to the national level. It also shows the faster growth in the number of deaths due to COVID-19 in Campos dos Goytacazes between the 21^st^ and 31^st^ E.W. compared to other municipalities.

The distribution of deaths by COVID-19 in the different age groups and by sex is shown in figure 3. The same general pattern is observed in all locations, with deaths concentrated in the central age groups (50 to 89 years), with male predominance in general, and few deaths at the extremes (children and young people, and the elderly over 90 years). Brazil and Rio de Janeiro have very similar distributions, whereas the municipalities analyzed show greater variability. Cabo Frio has a more even distribution between the ages of 50 and 89, and Rio das Ostras a distribution particularly concentrated in the 60-year-old male range.

**Figure 3.**
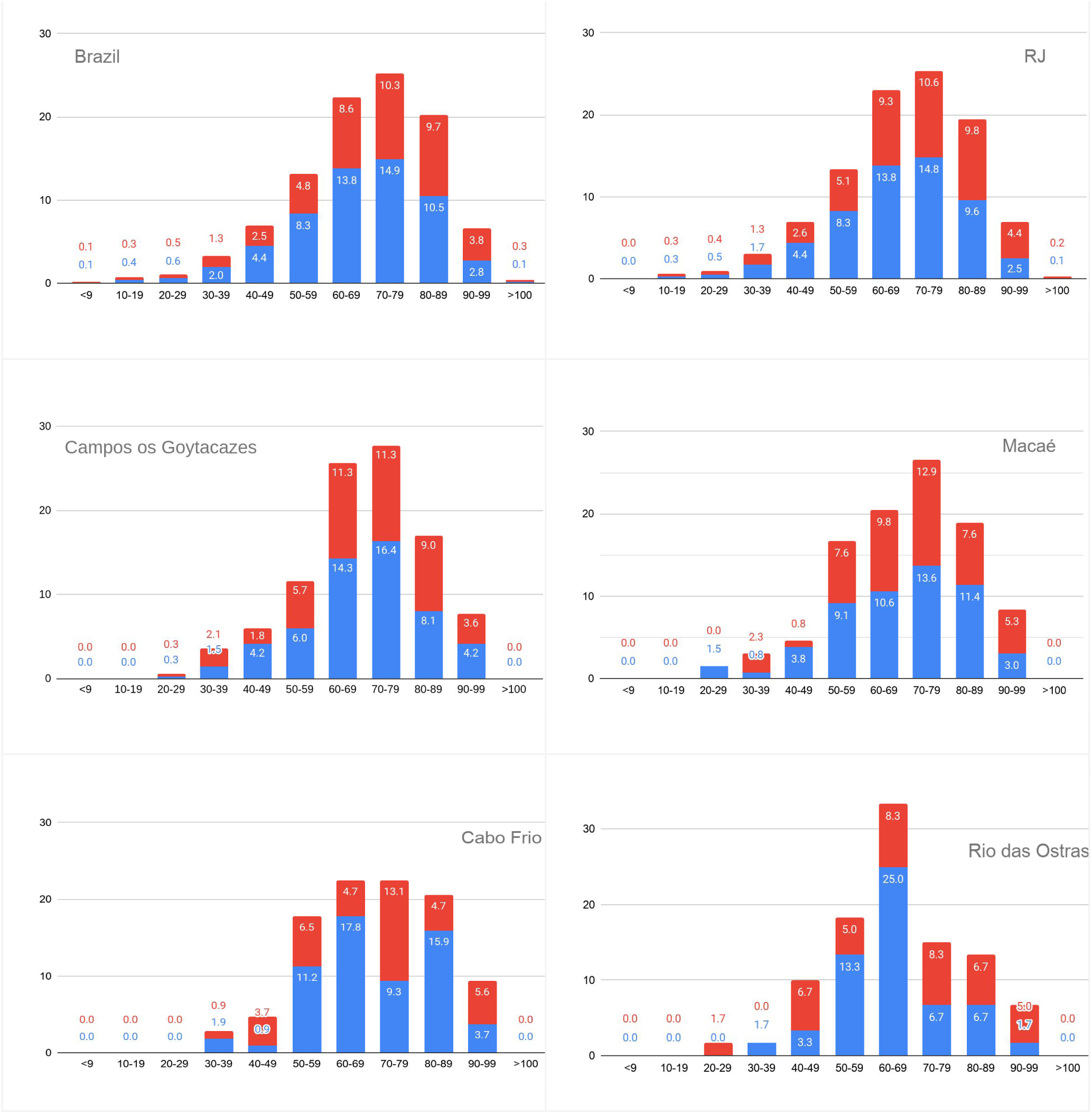
Percentage of deaths from COVID-19 by age group (in years) and sex (men in blue at the bottom of the bars, women in red at the top of the bars).

Population groups divided by age and sex can have very different populations, so it is interesting to calculate the mortality coefficient for each group, which allows for a more significant comparison. The difference in the outcome of the disease between the female and male populations is best observed in Table 1. The largest increase was found in Cabo Frio, where the mortality coefficient for COVID-19 was 63% higher for the male population compared to the female population. In turn, the lowest proportion was found in Macaé where the male population has a mortality rate 19% higher than that of women.

**Table 1.**
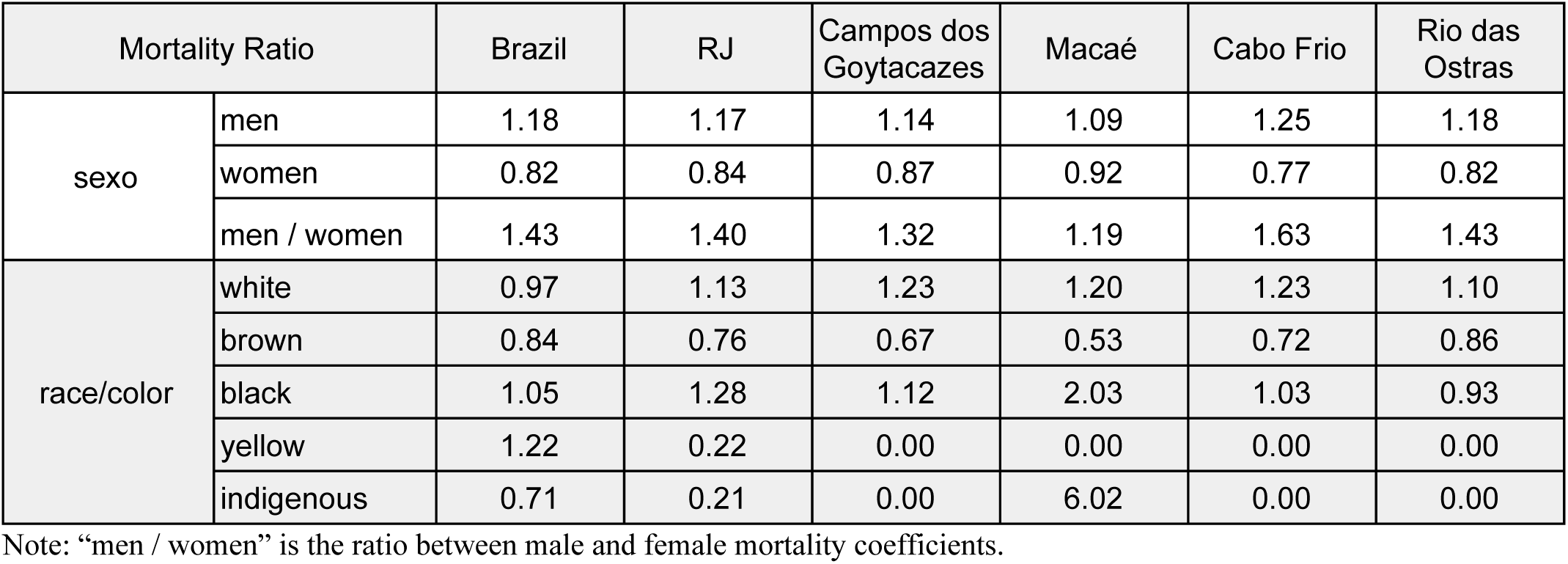
COVID-19 mortality compared between sex and race/color groups. Mortality ratios are presented (ratio of the group’s mortality coeficient to the mortality coeficient for the entire population of the localities).

| Mortality Ratio |  | Brazil | RJ | Campos dos Goytacazes | Macaé | Cabo Frio | Rio das Ostras |
| --- | --- | --- | --- | --- | --- | --- | --- |
| sexo | men | 1.18 | 1.17 | 1.14 | 1.09 | 1.25 | 1.18 |
|  | women | 0.82 | 0.84 | 0.87 | 0.92 | 0.77 | 0.82 |
|  | men / women | 1.43 | 1.40 | 1.32 | 1.19 | 1.63 | 1.43 |
| race/color | white | 0.97 | 1.13 | 1.23 | 1.20 | 1.23 | 1.10 |
|  | brown | 0.84 | 0.76 | 0.67 | 0.53 | 0.72 | 0.86 |
|  | black | 1.05 | 1.28 | 1.12 | 2.03 | 1.03 | 0.93 |
|  | yellow | 1.22 | 0.22 | 0.00 | 0.00 | 0.00 | 0.00 |
|  | indigenous | 0.71 | 0.21 | 0.00 | 6.02 | 0.00 | 0.00 |

Note: “men / women” is the ratio between male and female mortality coefficients.

The straight line around which the points are concentrated in figure 4 reveals an exponential dependence of the mortality coefficient of COVID-19 with age, regardless of location. Adjusting the function *y*(*x*) = *A* exp(α*x*), with *x* representing the average value in years in each age range, gives *A* = (4,55 ± 0.68) deaths per million and α = (0.0887 ± 0,0030) years^-1^ for Brazil (age group over 100 excluded from adjustment). All locations exhibit the same type of behavior, well described by an exponential up to the age of less than 100 years. It is possible that the accuracy of the age coefficient of mortality values is impaired by the use of the 2010 census age pyramid, as the age distribution of the population in different locations may have changed.

**Figure 4.**
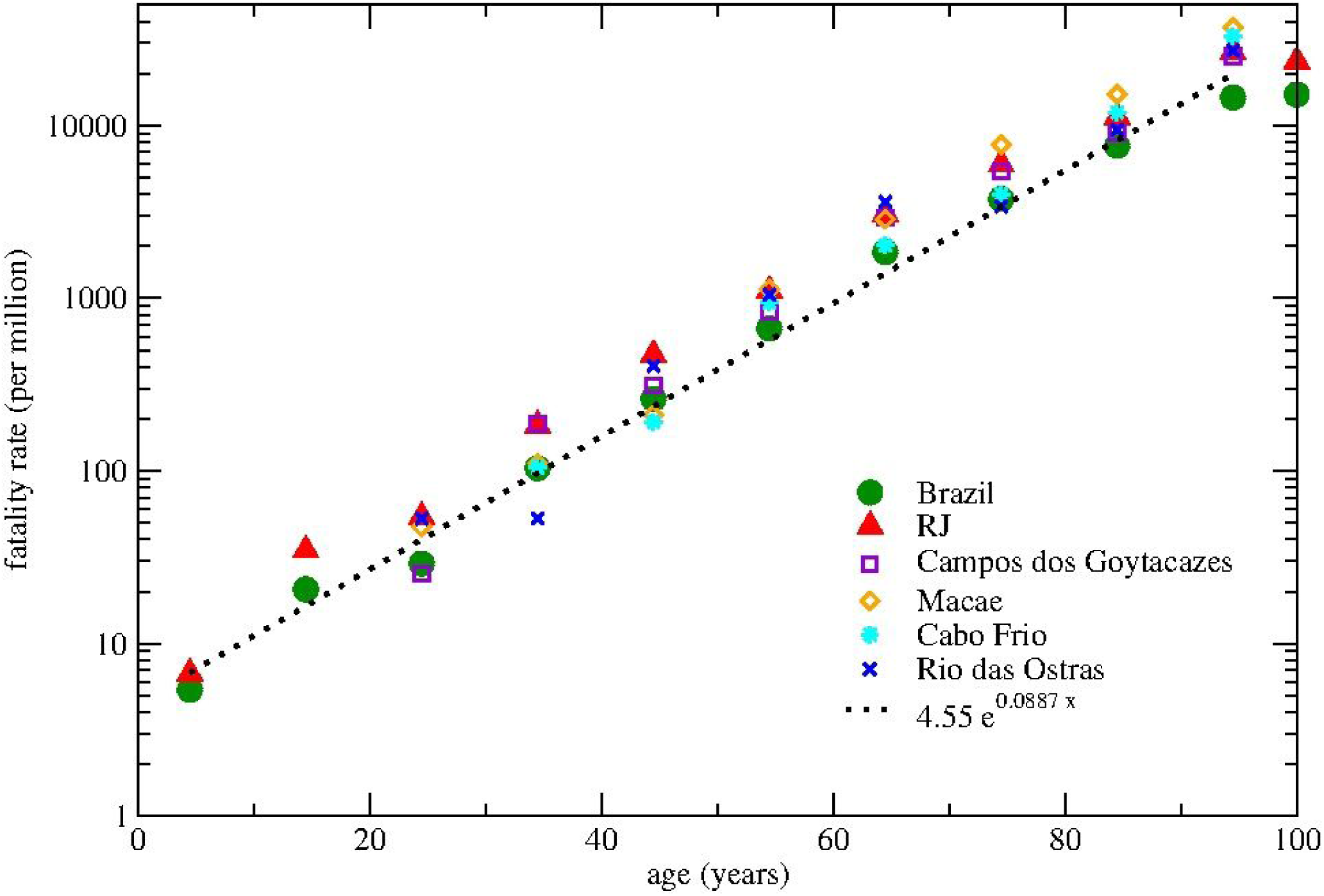
COVID-19 mortality coefficient (on a log scale) by age group.

Table 1 shows the ratio of the COVID-19 mortality coefficient for specific groups (sex and race/color) to the general coefficient in a given location (which we define as the mortality ratio), thus allowing the comparison of mortality between groups and locations. Race evaluation is relevant for a better understanding of the behavior of a disease in different individuals. The “yellow” and “indigenous” race/color groups, as they are of low representativeness in the evaluated cities, were not discussed. It is noted, when analyzing the mortality ratios of the “white”, “brown” and “black” groups in the localities, that the “brown” population presented the lowest value in all the localities. The “black” group has the highest mortality rate in Brazil, in the state of Rio de Janeiro and in Macaé. While the “white” group had the highest mortality rate in Campos dos Goytacazes, Cabo Frio and Rio das Ostras.

Table 2 provides a general picture of various causes of death, comparing the mortality rate of each cause in the period considered in the years 2019 and 2020, with which we perceive the direct and indirect impacts of COVID-19. In 2020 COVID-19 represents 18% of deaths in Brazil, 22% in the State of Rio de Janeiro, 18% in Campos dos Goytacazes, 21% in Macaé, 14% in Cabo Frio and 17% in Rio das Ostras, being major cause of death, after “other”, among those listed in the table.

**Table 2.** Comparative chart of *Causa Mortis* (mortality coefficient per million inhabitants) in the period from Mar/15 to Aug/01 (20 weeks). “Var.%” Is the percentage change between 2019 and 2020. Mortality at home is also shown.

|  | Brazil |  |  | Rio de Janeiro |  |  | Campos |  |  | Macaé |  |  | Cabo Frio |  |  | Rio das Ostras |  |  |
| --- | --- | --- | --- | --- | --- | --- | --- | --- | --- | --- | --- | --- | --- | --- | --- | --- | --- | --- |
| *causa mortis* | 2019 | 2020 | Var. % | 2019 | 2020 | Var. % | 2019 | 2020 | Var. % | 2019 | 2020 | Var. % | 2019 | 2020 | Var. % | 2019 | 2020 | Var. % |
| **COVID-19** | **0** | **524** |  | **0** | **868** |  | **0** | **673** |  | **0** | **505** |  | **0** | **464** |  | **0** | **387** |  |
| respiratory failure | 188 | 171 | -9 | 192 | 206 | 7 | 160 | 168 | 5 | 149 | 103 | -31 | 208 | 182 | -13 | 135 | 187 | 38 |
| pneumonia | 445 | 307 | -31 | 540 | 462 | -14 | 818 | 640 | -22 | 413 | 298 | -28 | 399 | 399 | 0 | 342 | 245 | -28 |
| septicemia | 333 | 274 | -18 | 500 | 391 | -22 | 516 | 354 | -31 | 444 | 252 | -43 | 321 | 291 | -9 | 322 | 226 | -30 |
| SARS | 4 | 56 | 1440 | 2 | 72 | 3015 | 0 | 22 |  | 4 | 19 | 400 | 0 | 26 |  | 0 | 13 |  |
| Stroke | 219 | 204 | -7 | 200 | 199 | 0 | 309 | 303 | -2 | 214 | 141 | -34 | 139 | 156 | 13 | 129 | 174 | 35 |
| heart attack | 222 | 186 | -16 | 283 | 255 | -10 | 194 | 256 | 32 | 145 | 122 | -16 | 260 | 300 | 15 | 219 | 200 | -9 |
| unspecified cardiovascular causes | 158 | 202 | 27 | 237 | 275 | 16 | 164 | 215 | 31 | 115 | 176 | 53 | 178 | 265 | 49 | 148 | 129 | -13 |
| indeterminate | 14 | 20 | 36 | 2 | 40 | 1653 | 25 | 27 | 8 | 0 | 50 |  | 26 | 95 | 267 | 0 | 0 |  |
| other | 998 | 968 | -3 | 1137 | 1118 | -2 | 1274 | 982 | -23 | 654 | 750 | 15 | 1046 | 1024 | -2 | 664 | 689 | 4 |
| **total** | **2586** | **2912** | **13** | **3094** | **3887** | **26** | **3461** | **3641** | **5** | **2138** | **2417** | **13** | **2578** | **3203** | **24** | **1959** | **2249** | **15** |
| deaths at home | 509 | 645 | 27 | 408 | 576 | 41 | 458 | 558 | 22 | 237 | 428 | 81 | 347 | 512 | 48 | 316 | 367 | 16 |

Note: data extracted on 08/27/2020

A measure of the global impact is the total excess of deaths from 2020 compared to 2019, which in Table 2 is expressed by the percentage variation of total deaths, which ranges from 5% for Campos dos Goytacazes to 24% for Cabo Frio at the municipal level, 26% in the state of Rio de Janeiro and 13% nationally. The number of deaths from COVID-19 is greater than the total excess of deaths in 2020 compared to 2019 in all locations, which means a reduction in total mortality from all causes other than COVID-19, except Cabo Frio, where the excess mortality was higher than the mortality due to COVID-19. In fact, some of the causes of death examined show a significant reduction in 2020 compared to 2019 in almost all locations, for example pneumonia and septicemia. Other causes show a fall or elevation depending on the location, such as respiratory failure, stroke and heart attack. On the other hand, Severe Acute Respiratory Syndrome (SARS) has grown significantly in all locations. Non-specific cardiovascular causes and undetermined cause also increased in all locations, with the exception of Rio das Ostras. The item “other” (causes of death), which includes other diseases, such as cancer, and also violent deaths, remained stable nationally in the state, in Cabo Frio and Rio das Ostras, but decreased significantly in Campos dos Goytacazes and grew in Macaé.

The mortality from pneumonia in Campos dos Goytacazes is noteworthy, something like twice that observed nationally. In Cabo Frio, there is a high mortality due to infarction, unspecified cardiovascular causes and undetermined cause, and it is surprising and worthy of investigation that all cardiovascular and undetermined causes of death had such a significant increase in mortality from 2019 to 2020. Macaé stands out for its low mortality due to respiratory failure, stroke and infarction. In Rio das Ostras, the location with the lowest mortality rates due to COVID-19 and total, there is low mortality due to SARS, unspecified cardiovascular causes and undetermined cause. The number of deaths at home increased significantly in all locations between 2019 and 2020, as shown in Table 2, with Macaé standing out with an 81% increase.

It is important to note that when calculating quantities such as the mortality coefficient, it is possible that there is some deviation from the real value, as deaths that occur in a given location do not take into account that people may die in other locations than in their hometown. From the CR data we can obtain the number of deaths by COVID-19 of non-residents in a given municipality (9.8% in the capital, 13.4% in Campos dos Goytacazes, 4.5% in Macaé, 11.2% in Cabo Frio and 38.3% in Rio das Ostras), but not the number of residents of a municipality who died outside it. Therefore, it was not possible to make these corrections in the mortality coefficients presented here.

## 4. Discussion

The monitoring of death indicators is a strategy recommended by the World Health Organization to assess the direct and indirect effects of the COVID-19 pandemic in countries^11^. The number of deaths from natural causes in Brazil has traditionally increased slightly every year due to the aging of the population. However, what we found was that the pandemic caused by SARS-CoV-2 had a major impact on mortality in the locations analyzed, from the municipal to the national level, not only directly through deaths from COVID-19, but also indirectly, affecting statistics on various causes of death and being associated with an excess of overall mortality compared to the previous year^12^.

We found that the pandemic had a later evolution and less reach in the analyzed municipalities than in the state as a whole, whose measures are dominated by the metropolitan region of the capital. However, the municipalities studied do not differ much from the national average regarding the evolution and accumulation of fatalities by COVID-19 in terms proportional to their populations, with the largest municipality (Campos dos Goytacazes) presenting the worst picture in terms of growth and coefficient of mortality, 51% above the national average, but still below the state average, which is almost twice the national average.

We also emphasize that the municipalities studied and the state of Rio de Janeiro maintain, after a phase of maximum, weekly mortality rates due to COVID-19 at levels close to the national level at the end of the period studied (which has not changed in the following E.W.). This may explain the maintenance of the national mortality rate at a practically constant level for such a long time, differing from the pattern of other countries affected by the pandemic. At the same time, if the mortality rate at the end of the period considered (with deaths continuing to accumulate at a significant rate later) for the state of Rio de Janeiro was taken as a measure of the potential reach of the pandemic for other locations municipally and nationally, this would indicate a very serious situation, projecting an accumulation of deaths that could be double that already observed at the end of the 31^st^ W.E. for some of these municipalities and for the country. The state of Rio de Janeiro is also a concern if we consider another analysis: from the beginning of the pandemic (12^th^ E.W.) to the peak of the mortality curve (19^th^ E.W.) the accumulated number of deaths is one third of the total accumulated deaths up to the 31^st^ E.W., or in other words, the “descent of the mortality curve” phase has a considerably greater integral than the “rise of the curve” phase, since the “descent” is slower than the “rise”, which also projects a potential number of deaths very large in the studied locations, having as reference the one already observed at the end of the 31^th^ E.W. Measures to relax control over the pandemic should be taken with caution.

We obtained results on COVID-19 mortality for specific groups: by age group, sex and race/color, however, mortality can depend both on the exposure to SARS-CoV-2 and on the severity of the resulting infection within the different population groups. Surveys on the prevalence of SARS-CoV-2 antibodies may help to differentiate between the two possibilities (exposure and severity of infection). The EPICOVID19-BR study, of national reach, indicated that: (i) there is no significant difference in prevalence between age groups and sex, but (ii) there is a significant difference (considering the confidence intervals) for the prevalence between race groups/color (the white, black and brown groups are increasingly prevalent)^13^.

Our results on the distribution of deaths by COVID-19 by age groups reveal that, when considering the population size of the respective groups, that is, based on the mortality coefficient for each age group, the infection is particularly lethal for seniors. So, considering that exposure to the virus is similar between age groups, it is concluded that the severity and, consequently, the fatal outcome of the disease, is higher in more advanced age groups. In this sense, it is believed that these patients have a higher proportion of comorbidities than younger age groups^2^. The lethality coefficient of COVID-19 (deaths from infected) is strongly dependent on age, which requires even stricter public and private care recommendations for the elderly.

We also found, as in international studies^14^, that male mortality due to COVID-19 is higher than that of women. As exposure to the virus appears to be the same among men and women, it is concluded that the infection is more severe in men. Socioeconomic and cultural factors related to gender have been pointed out to explain the higher male mortality^15^, but there are also hypotheses of genetic and immunological origin^16, 17, 18^.

We believe that our results and analyzes on the race/color factor should be considered incipient and encourage further studies^19,20^. The analysis of COVID-19 mortality among race/color groups is more complex, as this population division is more subjective and performed differently in different situations. In census and prevalence surveys, self-identification predominates, but in medical records and death certificates the classification can be made by a family member or health professional. Another complicating factor is that population groups divided by race/color may have different internal distributions by sex and age. As we have already concluded that fatality depends strongly on age and also on sex, by not isolating these factors in an analysis by race/color one may incur in bias.

Several causes of death, such as respiratory failure, pneumonia, septicemia, stroke and infarction had a reduction in the national level when compared to 2019. In the state of Rio de Janeiro and in the analyzed municipalities, some of the causes have a reduction, others increase, forming a complex general situation. It is possible to speculate that the pandemic may have indirect effects in reducing mortality due to some of these causes as a result of social distancing and extra hygiene measures practiced by individuals that also saves them from exposure to other pathogens and risk situations of other diseases and even accidents. On the other hand, it is to be expected that individuals with health conditions conducive to the worsening of certain diseases (such as pneumonia or heart disease, for example), if exposed to SARS-CoV-2, would be much more likely to develop a severe condition of COVID-19. Thus, deaths that in the absence of SARS-CoV-2 could occur due to other diseases, end up occurring when people contract COVID-19.

Deaths due to SARS, nonspecific cardiovascular causes and undetermined causes have grown nationally, compared to the year 2019, possibly indicating underreporting of mortality by COVID-19 itself or also an overload of the health system and health professionals, making it difficult to accurately identify the cause mortis. The increase, in all studied locations, of deaths occurring at home corroborates this hypothesis.

The COVID-19 pandemic directly and indirectly impacts the death toll of municipalities, state and country, which our study helped to know and understand, as well as raising new questions. These results should encourage authorities to draw on the growing scientific knowledge about the pandemic for decision-making and protection of those who are most vulnerable, avoiding deaths and overloading the health system due to the large number of people at greatest risk of developing severe disease in case of COVID-19 infection.

## Data Availability

All data are publicly available.

## Acknowledgment

To Franci de Oliveira Barros, biologist at the Municipality of Macaé and a coder of death in the use of the CID-10 by the State Secretary of Health of Rio de Janeiro, for the discussions and clarifications about the database.

